# Breaking The Pain-Stiffness Cycle-Supraclavicular Catheter Facilitated Rehabilitation Of Post-Surgical Elbow stiffness-A Retrospective Observational Study

**DOI:** 10.64898/2026.06.14.26355590

**Authors:** Sunandha Mukundan, Vijaya Gandipan Arulanandham

**Author notes:** Address correspondence to: Dr Sunandha Mukundan, DNB, Department of Anesthesiology, MIOT International Hospital, Mount Poonamalee High Road, Manapakkam, Chennai 600089, India. Funding: The authors have no sources of funding to declare for this manuscript. AI Disclosure: The authors declare the following use of Artificial Intelligence in the creation of the manuscript: Artificial intelligence tools were used for grammar checking, sentence refinement and clarity enhancement during the manuscript preparation. This AI use was limited to language enhancement and did not influence scientific content, methodology or conclusions.

## Abstract

**Background:** Post-traumatic elbow stiffness is a recognised complication following orthopaedic trauma surgery, occurring in 10-15% of trauma patients sustaining injuries. Pain remains the primary barrier to physiotherapy compliance, with surgical arthrolysis carrying recurrence rates of up to 34%. The supraclavicular brachial plexus block, referred to as the ‘spinal of the arm’, provides anaesthesia and analgesia to the entire upper limb below the shoulder. A structured non-surgical approach combining continuous catheter analgesia with timed rehabilitation was identified as an unmet need in this patient group.

**Methods:** A single-centre retrospective observational study was conducted on data of patients treated for post-surgical upper limb stiffness between January 2022 and April 2026. Of 30 patients identified, 28 with elbow involvement formed the primary analysis group following exclusion of 2 patients with isolated wrist stiffness and complex regional pain syndrome. Ultrasound-guided supraclavicular brachial plexus catheters were inserted using the Contiplex system. Patients received 0.5% Bupivacaine (10-15ml) for initial blockade, followed by daily top-up doses of 0.2% Ropivacaine(20ml) given 30 minutes prior to structured physiotherapy and CPM sessions for up to 5 days. The primary outcome was change in arc of elbow motion in degrees, measured by the attending orthopaedic consultant using standard goniometry.

**Results:** Complete pre- and post-intervention data were available for all 28 patients. Mean pre-intervention arc of elbow motion was 39.1°(SD+/-23.2°), improving to 104.2°(SD+/-30.0°) post-intervention. Mean improvement was 65.1°(SD+/-30.6°; 95% CI 53.8° to 76.4°; range 10°-140°; paired t-test t=-11.27, p<0.0001). Mean catheter duration was 5.2 days (SD+/-1.3). Five patients (17.9%) experienced mechanical catheter complications-3 dislodgements, 1 kinking and 1 block failure-with no episodes of Local Anaesthetic Systemic toxicity, infection or neurological deficit.

**Conclusion:** Continuous supraclavicular brachial plexus catheter analgesia represents a promising, minimally invasive rehabilitation tool for post-surgical elbow stiffness — achieving statistically and clinically meaningful Range of Motion (ROM) improvement through targeted regional analgesia, without the need for surgical intervention. These findings support prospective evaluation of this protocol as a primary non-surgical rehabilitation strategy.

## Introduction

Post-traumatic elbow stiffness is a recognised complication following elbow injury, with an occurrence rate of 10%–15% among trauma patients. A 50% reduction in elbow range of motion can lead to an 80 % decline in upper limb functionality, significantly impairing work performance and social participation.^1^

Following trauma, surgery, inflammation, pain and immobilization often lead to decreased range of movement, secondary hyperalgesia and delayed recovery. Any loss or restriction of the elbow movement due to elbow stiffness can cause substantial disability. Current conservative management strategies include physiotherapy, splinting and Continuous Passive Motion (CPM) -all effective only when the patient can tolerate mobilisation. Pain remains the primary barrier limiting compliance and outcome. When conservative measures fail, surgical arthrolysis remains the next option - however recurrence rates of up to 34% and associated surgical morbidity limit its universal applicability. ^2^

The supraclavicular brachial plexus block commonly referred as the ‘spinal of the arm’ provides anaesthesia and analgesia to entire upper limb below shoulder covering elbow, forearm, wrist and hand. ^3^ An ultrasound guided placement of a continuous catheter can provide continuous analgesia for days. ^4^

While continuous peripheral nerve blocks have demonstrated benefit in postoperative analgesia and rehabilitation following elbow arthrolysis, their role as a primary rehabilitation strategy for established post-traumatic upper limb stiffness remains largely unexplored. Furthermore, a structured protocol combining supraclavicular catheter analgesia with timed CPM and physiotherapy has not been previously described.

A strategy that eliminates pain during rehabilitation, enabling effective physiotherapy and CPM without surgical intervention, represents an unmet gap in management of the stiffness.

This retrospective study aimed to analyse the efficacy and safety of continuous supraclavicular brachial plexus catheter analgesia as a primary non-surgical rehabilitation strategy in patients with established post-surgical elbow stiffness - observing the Range of Movement (ROM) improvement, catheter outcomes and complications as the primary outcome.

## METHODS

### Study Design and Setting

A single-centre retrospective observational study was conducted at MIOT International Hospitals, Chennai, India. Data were collected from patients treated for post-surgical upper limb stiffness between January 2022 and April 2026. Ethical approval was obtained from the Institutional Ethics Committee, as a retrospective observational study, individual patient consent for data analysis was waived by the committee. The study was conducted in accordance with the Declaration of Helsinki.

### Patient Selection

Records of all patients who underwent supraclavicular brachial plexus catheter intervention for post-surgical elbow stiffness during the study period were retrospectively reviewed. Cases were eligible for analysis if the patient was aged 18– 80 years, had documented post-surgical elbow stiffness of minimum 6 weeks duration and had complete pre- and post-intervention range of motion data available.

Cases were excluded from analysis if documentation was incomplete, if the patient had isolated wrist or finger stiffness without elbow involvement, or if the catheter had been inserted for an indication other than rehabilitation of joint stiffness, like Complex Regional Pain Syndrome (CRPS).

As a retrospective study, selection bias was minimised by including all consecutive eligible records identified during the study period without outcome-based exclusion.

The flow of records through the study is illustrated in Figure 1.

**Figure 1.**
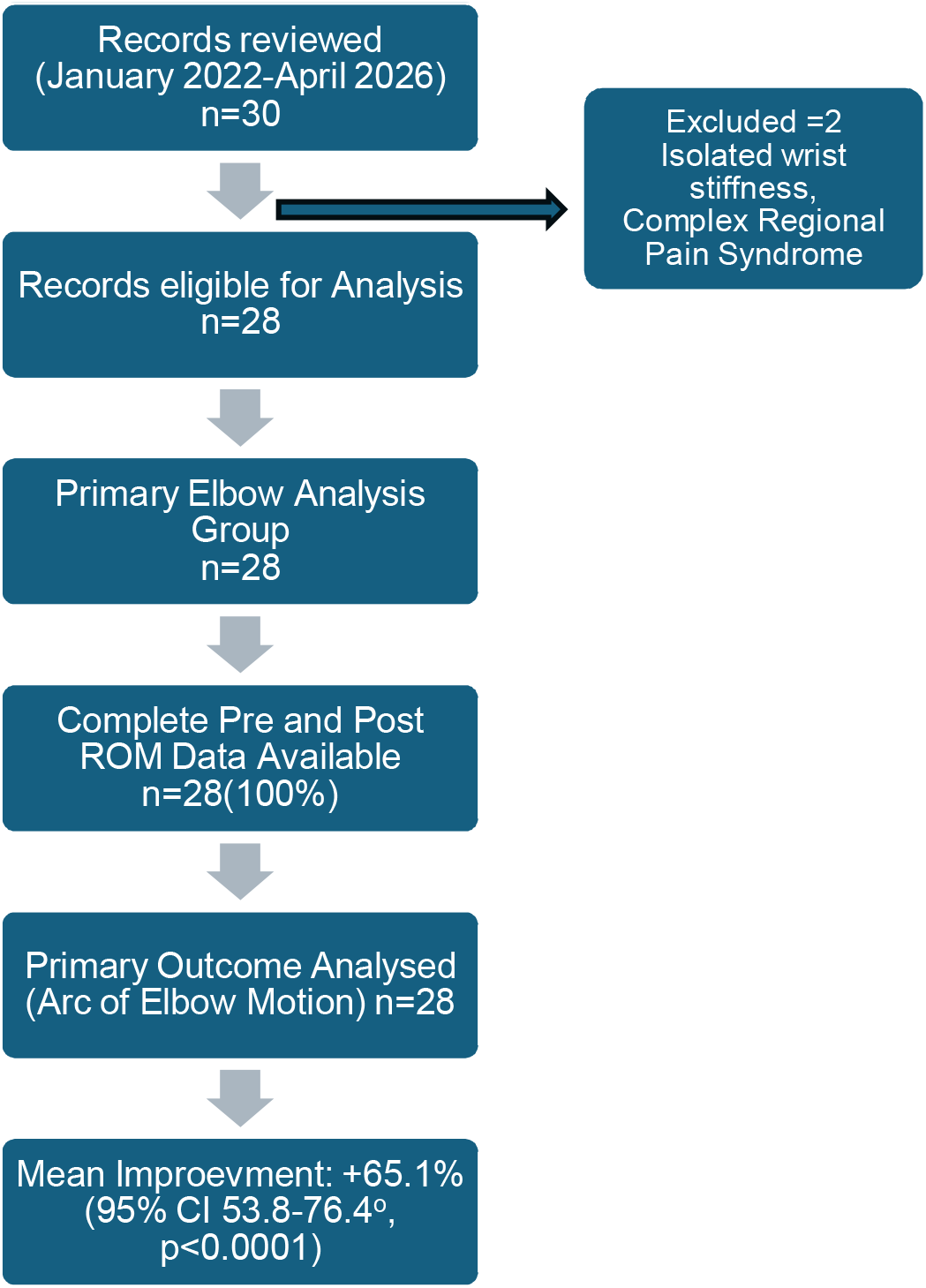
Flow diagram of records

### Intervention

The intervention protocol followed at the institution during the study period was retrospectively documented as follows. Ultrasound-guided in-plane supraclavicular brachial plexus catheter placement was performed in all cases using the 20G Contiplex stimulating catheter system (B.Braun). Intravenous sedation was administered with Midazolam 1mg and Fentanyl 50mcg. Following local infiltration with 2% Lignocaine 2ml, the needle was advanced in-plane with linear high frequency probe under real-time ultrasound guidance (Sonosite), the catheter threaded into the target periplexus area, distribution confirmed with cold saline, and initial plexus blockade achieved with 0.5% Bupivacaine 10–15ml. A sterile dressing was applied and the catheter secured with a sterile connector.

On Day 0, the first mobilisation session was conducted 30 minutes following initial blockade. From Day 1 onwards, 20ml of 0.2% Ropivacaine was administered once daily through the catheter 30 minutes prior to the morning mobilisation session, as per the institutional protocol. Structured physiotherapy and Continuous Passive Motion (CPM) were conducted twice daily for up to 5 days, incorporating passive and active techniques progressively. The catheter was removed on completion of the 5-day protocol or earlier in the event of displacement, kinking or block failure. In cases of premature catheter removal, rehabilitation was continued with supplemental systemic analgesia.

### Outcome Measures

The primary outcome was change in arc of elbow motion in degrees ROM, calculated as the difference between pre- and post-intervention measurements. Range of motion was measured by the attending orthopaedic consultant using standard goniometry before catheter insertion and after mobilisation. Secondary outcomes included catheter duration, complication rates and the degree of functional restoration.

### Statistical Analysis

Data were analysed using paired t-test to compare pre- and post-intervention arc of motion. Results are presented as mean ± standard deviation (SD) with 95% confidence intervals (CI). A p value of less than 0.05 was considered statistically significant. Statistical analysis was performed using Python (version 3.12, SciPy library). As a retrospective observational study, formal sample size calculation was not performed; all consecutive patients meeting inclusion criteria during the study period were included.

## RESULTS

Data from 30 patients who had undergone continuous supraclavicular brachial plexus catheter intervention for post-surgical elbow stiffness were analysed (Figure 1). Two records were excluded — one with complex regional pain syndrome and one with isolated wrist stiffness-as elbow range of motion was not assessable. The remaining 28 patients formed the primary elbow analysis group with complete pre- and post-intervention data available for all 28 patients.

### Demographics

Demographic characteristics are summarised in Table 1. The cohort comprised 20 males and 8 females with a mean age of 43.8 years (range 26–67 years). The mean duration of upper limb stiffness prior to intervention was 4.1 months (range 6 weeks to 20 months). Two patients presented with prolonged stiffness exceeding one year.

**Table 1.** Patient Demographics.

| Parameter | Value |
| --- | --- |
| Total studied | 30-2=28 |
| Male / Female | 20 / 8 |
| Mean age | 43.8 years (range 26–67) |
| Mean stiffness duration | 4.1 months (range 6 weeks – 20 months) |
| Mean catheter duration | 5.2 days (SD $\pm 1.3$, range 2–8) |

### Primary Outcome — Elbow Range of Motion (n=28)

Complete pre- and post-intervention range of motion data were available for all 28 patients. The mean pre-intervention arc of elbow motion was 39.1° (SD ±23.2°) and mean post-intervention arc was 104.2° (SD ±30.0°), representing a mean improvement of 65.1° (SD ±30.6°; 95% CI 53.8° to 76.4°; range 10° to 140°). This improvement was statistically significant on paired t-test (t=−11.27, p<0.0001). Results are summarised in Table 2.

**Table 2.**
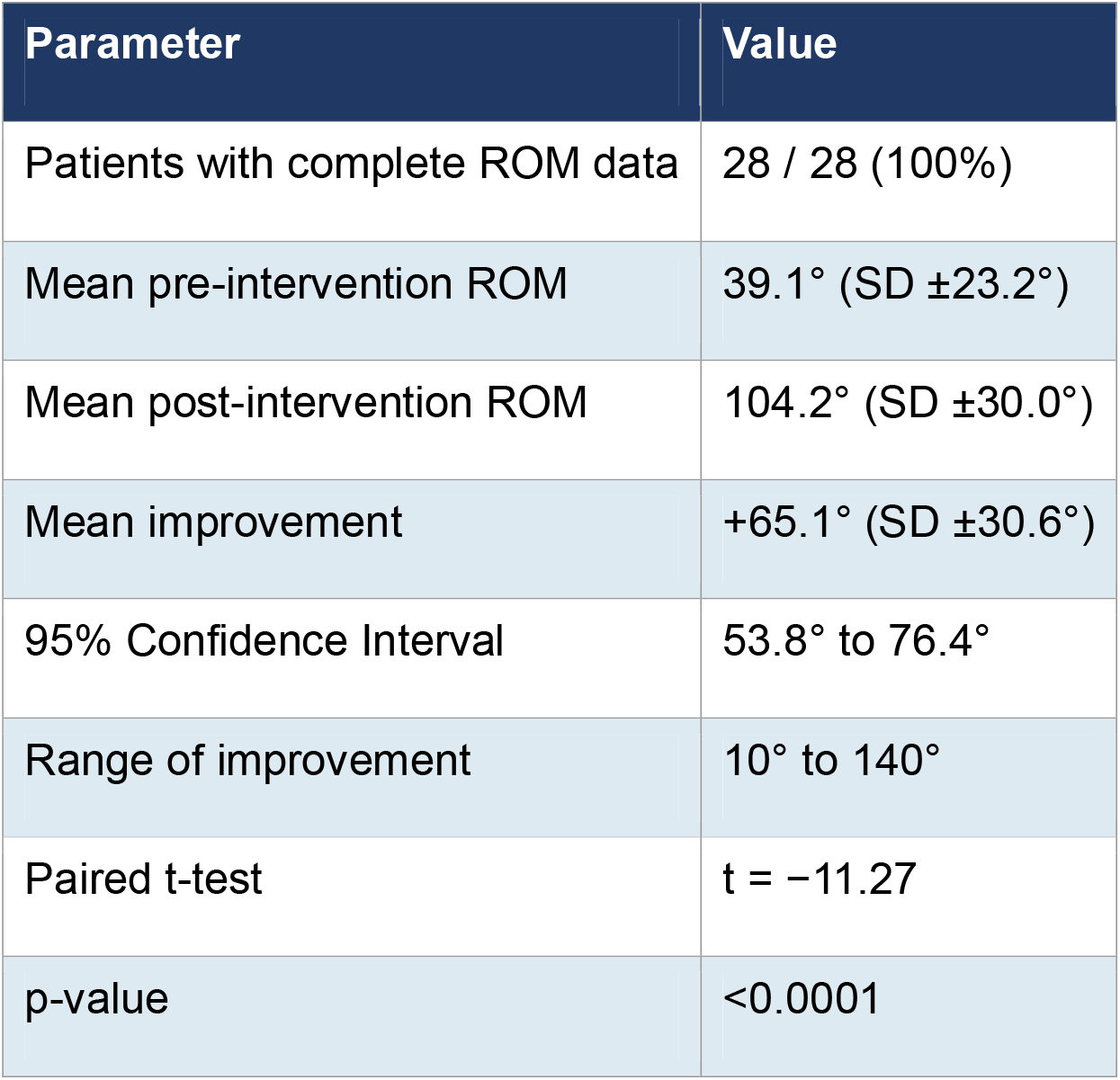
Primary Elbow Group — Range of Motion Outcomes.

### Complications

Five patients (17.9%) experienced catheter-related mechanical complications resulting in premature catheter removal - three dislodgements (10.7%), one catheter kinking (3.6%) and one block failure (3.6%). In these cases, the rehabilitation protocol was continued with supplemental systemic analgesia and transdermal Buprenorphine patch. No adverse events including allergic reactions, local anaesthetic systemic toxicity (LAST), infection or neurological deficits were documented. Complications are summarised in Table 3.

**Table 3.** Catheter-Related Complications.

| Complication | Number (n) | Percentage (%) |
| --- | --- | --- |
| Catheter dislodgement | 3 | 10.7 |
| Catheter kinking | 1 | 3.6 |
| Block failure | 1 | 3.6 |
| Total | 5 | 17.9 |

## DISCUSSION

This retrospective observational study demonstrated that continuous supraclavicular brachial plexus catheter analgesia, combined with timed CPM and physiotherapy, achieved a mean arc of elbow motion improvement of 65.1° (95% CI 53.8° to 76.4°, p<0.0001) in 28 patients with established post-surgical upper limb stiffness. All patients had failed standard postoperative physiotherapy prior to intervention, making catheter analgesia the sole new variable introduced. The observed improvement therefore suggests that pain elimination during mobilisation was the critical facilitating factor enabling effective rehabilitation in the observed outcomes.

Further support for catheter utility was observed in the complication subgroup. Patients experiencing premature catheter removal due to mechanical failure demonstrated comparatively reduced range of motion improvement —the patient whose catheter dislodged on Day 4, achieved only +10°, and whose catheter failed on Day 2, achieved only +25° — compared to a mean improvement of 65.1° in the overall cohort. While these observations are based on small numbers and confounding factors cannot be excluded, the pattern suggests that sustained catheter analgesia may be important for optimising rehabilitation outcomes.

There is a notable gap in literature on the use of peripheral nerve block catheters as a primary non-surgical rehabilitation strategy for established post-traumatic elbow stiffness. Babasiz et al. reported significant range of motion improvement following peripheral nerve block catheter after surgical elbow arthrolysis.^5^ The present study demonstrated comparable gains without surgical intervention and despite more severe baseline stiffness, suggesting the catheter plays a significant rehabilitative role independent of surgical release. Aguirre et al. established the role of continuous peripheral nerve block catheters in perioperative and chronic pain settings ^6^; this study extends that application to a novel indication of post-surgical upper limb stiffness rehabilitation. Jiménez Galán et al. utilised interscalene catheter for shoulder stiffness rehabilitation over 72 hours ^7^; a longer 5-day protocol was employed in this study for the anatomically distinct elbow joint, demonstrating meaningful sustained gains. Extensive literature supports femoral nerve block facilitating knee rehabilitation^8^ ; the present study provides an upper limb equivalent, establishing nerve block facilitated rehabilitation as a translatable concept across joints and anatomical regions. Collectively, these findings position continuous supraclavicular catheter analgesia as a clinically relevant, minimally invasive rehabilitation tool bridging the gap between failed conservative management and surgical intervention.

Post-traumatic elbow stiffness is frequently associated with fibrotic capsular contracture, necessitating effective analgesia during forceful mobilisation. The institutional protocol utilised Bupivacaine for initial blockade, exploiting its dense motor and sensory blockade properties, followed by ropivacaine top-up doses — well documented to provide preferential sensory blockade at lower concentrations while preserving motor function. ^9^ This pharmacological approach as per the institutional protocol, provided a structural analgesic framework where the progressive rehabilitation was conducted.

## LIMITATIONS

This study has several limitations. As a single-centre retrospective observational study with no control group, causal conclusions cannot be drawn. The use of paired pre- and post-intervention measurements provided a reasonable internal comparator, but absence of a physiotherapy-only control group limits attribution of improvement solely to the catheter. Range of motion was assessed by the treating orthopaedic consultant rather than an independent blinded assessor, introducing potential measurement bias. The heterogeneity of diagnoses and stiffness durations introduces variability that may influence outcomes. Variable catheter duration resulting from mechanical complications may have affected rehabilitation completeness in affected cases. Absence of structured long-term follow-up limits evaluation of durability of gains achieved.

## Conclusions

Continuous supraclavicular brachial plexus catheter analgesia, combined with timed CPM and physiotherapy, achieved statistically and clinically significant elbow range of motion improvement in patients with established post-surgical upper limb stiffness The catheter facilitated rehabilitation by eliminating the pain barrier, enabling effective physiotherapy compliance. Future prospective randomised controlled trials with control groups and long-term follow-up are warranted to validate these findings and establish this protocol as a part of rehabilitation strategy.

## Data Availability

All data produced in the present study are available upon reasonable request to the corresponding author. Individual patient data are not publicly available to protect patient privacy; anonymised summary data are contained in the manuscript.

## Acknowledgement

The authors wish to acknowledge Dr Krishna Shankar, Chief Anesthesiogist and Head of Trauma, Dr Ram prasad, Head of Department of Orthopaedics and Trauma for the clinical oversight, knowledge and guidance throughout the study. The authors also acknowledge Dr Prem C Sundar, Dr Akshara Purushothaman and Dr. Khalki Saravan kumar consultant anaesthesiologists for their contributions to catheter insertion and perioperative pain management. The anaesthesia technicians and nursing staff are acknowledged for their diligent catheter care patient monitoring and documentation throughout the inpatient rehabilitation programme.

